# Development of a Novel Medium Throughput Flow-Cytometry Based Micro-Neutralisation Test for SARS-CoV-2 with Applications in Clinical Vaccine Trials and Antibody Screening

**DOI:** 10.1101/2023.01.18.23284713

**Authors:** Sophie O’Reilly, Grace Kenny, Tamara Alrawahneh, Nathan Francois, Matthew Angeliadis, Valentin de Masson d’Autume, Alejandro Garcia Leon, Eoin R. Feeney, Obada Yousif, Aoife Cotter, Eoghan de Barra, Mary Horgan, Patrick WG Mallon, Virginie Gautier

## Abstract

Quantifying neutralising capacity of circulating SARS-COV-2 antibodies is critical in evaluating protective humoral immune responses generated post-infection/post-vaccination. Here we describe a novel medium-throughput flow cytometry-based micro-neutralisation test to evaluate Neutralising Antibody (NAb) responses against live SARS-CoV-2 Wild Type and Variants of Concern (VOC) in convalescent/vaccinated populations. Flow Cytometry-Based Micro-Neutralisation Test (Micro-NT) was performed in 96-well plates using clinical isolates WT-B, WT-B.177.18 and/or VOCs Beta and Omicron. Plasma samples (All Ireland Infectious Diseases (AIID) Cohort) were serially diluted (8 points, half-log) from 1/20 and pre-incubated with SARS-CoV-2 (1h, 37°C). Virus-plasma mixture were added onto VERO E6/VERO E6 TMPRSS2 cells for 18h. Percentage infected cells was analysed by automated flow cytometry following trypsinisation, fixation and SARS-CoV-2 Nucleoprotein intracellular staining. Half-maximal Neutralisation Titres (NT50) were determined using four-parameter logistic regression. Our assay was compared to Plaque Reduction Neutralisation Test (PRNT) and validated against WHO anti-SARS-CoV-2 Immunoglobulin Standards. Using WHO Standards with low, medium or high anti-SARS-CoV-2 IgG, both Micro-NT and PRNT achieved comparable NT50 values. Micro-NT was found to be highly reproducible (inter-assay CV of 11.64%). Screening 190 convalescent samples and 11 COVID-19 naive controls (AIID cohort) we demonstrated that Micro-NT has broad dynamic range differentiating NT50s <1/20 to >1/5000. We could also characterise immune-escape VOC observing up to 10-fold reduction in NT50 against SARS-CoV-2 Beta variant. Our flow cytometry-based Micro-NT is a robust and reliable assay to quantify NAb titres, and has been selected as an endpoint in clinical trials. It has higher throughput (96 well format versus 12 well) and reduced infection time (18h vs 48-96h) compared to the gold standard PRNT.

## Introduction

SARS-CoV-2 is the viral agent responsible for the Coronavirus Infectious Disease 2019 (COVID-19) pandemic (1). The disease was given pandemic status by the World Health Organisation in March 2020, and as of August 2022, there has been 600 million cases, and 6.5 million deaths (2).

Both T-cell and humoral immune responses are required for protection from COVID-19. Humoral immunity relies on B cell exposure to SARS-CoV-2 antigens, which triggers their proliferation into antibody secreting plasma cells (3). Following infection, antibodies are produced against SARS-CoV-2 viral proteins, predominantly the Spike (S) and the Nucleocapsid Protein (NP) (4, 5). Neutralising antibodies (NAbs) are a subset of SARS-CoV-2 antibodies that prevent viral entry, through either direct blocking of virus binding to the host cell receptor, or preventing conformational changes required for membranes fusion. SARS-CoV-2 NAbs target the S protein, making it the preferred COVID-19 vaccine candidate (6, 7).

The S protein is comprised of trimeric S1/S2 heterodimers. S1, harbouring an N-terminal domain and a Receptor Binding Domain (RBD), interacts with the host cell through binding of the RBD to the angiotensin-converting enzyme 2 (ACE-2) receptor. Following S1/S2 cleavage by host cell proteases including furin, S2’ cleavage by TMPRSS2 or Cathepsin-L mediates membrane fusion and cell entry (8). The most potent NAbs target the RBD as these directly compete with ACE-2 for binding. Mutations in this site are often associated with immune escape (9). Non-RBD sites are more evolutionarily conserved, so NAbs targeting these sites can often maintain efficacy against SARS-CoV-2 variants as well as display cross-reactivity with other sarbecoviruses (10).

An effective antibody response provides protection against COVID-19. ‘The Protective Neutralisation Classification Model’ described by (6) suggests that the protective neutralisation titre (reducing risk of infection by 50%) is 20% of the mean neutralisation titre of a convalescent cohort, which they have found to strongly predict protective immunity (against symptomatic disease) elicited by COVID-19 vaccine trials, while achieving a titre of only 3% of the mean is sufficient to reduce risk of severe disease by 50%. Not all S-targeting antibodies are neutralising. Post-infection or post-vaccination, a polyclonal antibody population is produced, targeting sites along the S protein (11). Some may only bind, but not have any neutralising capacity due to their site of action. Others may offer protection against Wild-type (WT) SARS-CoV-2, the strain against which the vaccine S is modelled, but not against immune-escape Variants of Concern (VOC), including Beta and Omicron, which have amino acid mutations in key antibody binding sites in the RBD (12). For this reason, an antibody titre, the measure of total anti-SARS-CoV-2 IgGs against a certain target present in a sample (13), is not sufficient to infer a protective immune response. Instead, the functional capacity of an antibody population can be determined using a neutralisation test.

Neutralisation tests, used to measure the capacity of a monoclonal antibody or plasma/serum to inhibit viral infection of susceptible cells, have proved valuable in elucidating SARS-CoV-2 antibody responses over time (14), in convalescent versus vaccinated individuals (15), and against SARS-CoV-2 VOCs (16) (17). This information is critical to forming effective public health strategies, from understanding when vaccine-induced protection wanes in different cohorts to devise booster strategies (18, 19), to identifying plasma donors for convalescent therapy (20, 21), to rapid identification of new VOCs that escape pre-existing immunity (22, 23).

The gold standard viral neutralisation assay is the Plaque Reduction Neutralisation Test (PRNT) (24). Here, live SARS-CoV-2 is co-incubated with serially diluted antibodies to facilitate neutralisation, and the virus is then used to infect a monolayer of cells. Over days, infected cells will display viral cytopathic effects (CPE) and die, leaving visible plaques in the monolayer that can be quantified to determine the reduction in infectious titre associated with the dilution factor of antibodies. However, this technique has several limitations which alternative assays have been developed to address. These include the large surface area for plaque formation requiring large wells, typically 6-or 12-well, which limits the throughput. The time to develop visual plaques can be up to 5 days for SARS-CoV-2. Furthermore, plaque counting is often done by eye, and even with automated software can be error-prone and subjective.

To address the low throughput, several assays have been adapted from PRNT, firstly facilitating neutralisation with a co-incubation of virus and antibodies, followed by determining the effectiveness of neutralisation through infection of susceptible cells. The key difference in these assays are the format (96-well plates) and the endpoints (Table 1). Rather than wait for visible plaques, micro-foci can be detected in the monolayer after only 18-hrs infection by staining of viral antigens (24, 25). Alternatively, recombinant SARS-CoV-2 virus expressing Green Fluorescent Protein can be detected directly without the need for further processing (25). Another approach is to detect viral genetic material in cell lysates through RT-qPCR following RNA extraction (26, 27) or to quantify replicating virus (viral load) by enzyme-linked immunosorbent assay (ELISA) (28). Other assays score the presence or absence of CPE under a microscope (29, 30) using a cell imager (31), or using colorimetric cellular dyes (29). All these assays have the advantage of being medium to high throughput, suitable for 96-well plates, and rapid compared to PRNT. However, they still have limitations. Foci-based assays can have difficulty separating spots that are close together, CPE is subjective and slow to develop, recombinant virus is time consuming to develop and validate to rapidly address new variants, and RT-qPCR is costly.

**Table 1:** Comparison of Micro-Neutralisation Tests. Micro-NTs are conducted in medium-throughput formats, typically 96-well plates. Following neutralisation of live SARS-CoV-2 by serial dilutions of plasma/serum, neutralisation capacity of each dilution is determined by corresponding infection levels compared to controls. The primary difference is the endpoint used to quantify infection, which may measure viral production directly, or viral cytopathic effects (CPE).

| Micro-Neutralisation Assays | Assay Endpoints | Infection time | Overlay required | Quantitative | Viral/Cellular Target | Reference |
| --- | --- | --- | --- | --- | --- | --- |
| Flow Cytometry Based Micro-NT | Flow Cytometry | 18-hrs | No | Yes | Intracellular SARS-CoV-2 NP | (This report) |
| ELISA-based virus neutralisation assay | Enzyme-linked immunosorbent assay (ELISA) | 24 hrs | No | Yes | Intracellular SARS-CoV-2 NP | (28) |
| qRT-PCR-based virus neutralization assay | RT-qPCR | 24 hrs | No | Yes | Intracellular SARS-CoV-2 genomic RNA | (26,27) |
| Microneutralization assay (MNA)/ Focus reduction neutralization test (FRNT) | Infected foci measured by computer-controlled imagers | 22-24 hrs | Yes | Yes | Cell-membrane associated SARS-CoV-2 Spike | (24, 25) |
| mNeonGreen-based FRNT (FRNT-mNG) | Fluorescent ELI-SPOT | 24-hrs | Yes | Yes | mNeonGreen expressing recombinant SARS-CoV-2 | (25) |
| Virus Neutralisation Test (VNT) | CPE observed under light microscope | 5 days | No | No | CPE | (30) |
| Colorimetric cytopathic effect-based microneutralization assay | Optical density of cellular lysate | 3 days | No | Yes | CPE proxy | (29) |
| High-content fluorescent live SARS-CoV-2 neutralization assay | Nuclei counts using IN CELL analyser | 3 days | No | Yes | CPE proxy | (31) |

One significant limitation of PRNT, is the use of live SARS-CoV-2. This means the assays must be carried out in Containment Level 3 facilities, by highly trained staff. Pseudovirus assays, where a viral backbone from vesicular stomatitis virus (VSV) or lentivirus, is engineered to express the SARS-CoV-2 S protein, have become popular choice as they only require Containment Level 2 facilities (32). However, they focus on the impact of S protein independently of other SARS-CoV-2 proteins present at the viral membrane and typically only measure viral entry, while live-virus neutralisation assays can monitor several rounds of replication. As the virus can spread cell-to-cell post-entry, it may evade neutralisation, thus explaining the lower neutralisation titres observed in such assays compared to pseudovirus assay (33). Surrogate Viral Neutralisation Tests or competitive immunoassays where antibodies prevent interaction between recombinant viral Spike protein and ACE-2 receptor *in vitro* have been proposed as a user-friendly method of inferring neutralisation capacity in the absence of virus, however they have shown poor correlation with PRNT (34).

Here we aimed to develop a novel neutralisation assay suitable for medium-throughput screening of SARS-CoV-2 antibodies. We chose a live virus assay, which most closely reflects *in vivo* neutralisation, as all viral proteins are present and complete. Instead of using a CPE-based approach, we used flow cytometry to quantify individual infected cells based on intracellular SARS-CoV-2 NP staining. This assay uses a 96-well plate format, for processing of up to 36-samples per day. It does not require washing the cells post-infection or adding an overlay, reducing hands-on processing time. The duration of infection is only 18-hrs allowing rapid turn-around of results, and the output is quantitative (% infected cells per well), less subjective than plaque or CPE-based assays.

This novel live SARS-CoV-2 flow-cytometry based micro-neutralisation test (Micro-NT) was calibrated and validated using WHO SARS-CoV-2 IgG standards and showed comparable neutralisation titres to PRNT. We show a high reproducibility of the assay and a broad dynamic range of neutralisation titres across a COVID-19 convalescent cohort. Finally, Micro-NT can immune escape of VOCs (Beta and Omicron-BA.5) compared to WT SARS-CoV-2 using the same plasma samples.

## Results

Here we aimed to develop a medium-throughput, rapid-turnaround live-virus Neutralisation Assay, to enable screening of large clinical cohorts for COVID-19 vaccine studies, COVID-19 trials for convalescent plasma therapy, or monoclonal antibody studies. The assay involves 5 steps (Figure 1). Firstly, serum from convalescent or vaccinated individuals is heat inactivated and serially diluted. Secondly, the antibody dilutions are co-incubated with live SARS-CoV-2 (viral neutralisation). Thirdly, the virus/antibody mixture is used to infect cells in culture. Next, cells are trypsinised and fixed, followed by intracellular staining and flow cytometry. Finally, the data is analysed to find the Neutralisation Titre resulting in a 50% inhibition of infection (NT50). The NT50 can then be used to easily compare the neutralisation capacities of human plasma between individuals, before or after vaccination, or against different SARS-CoV-2 variants.

**Figure 1:**
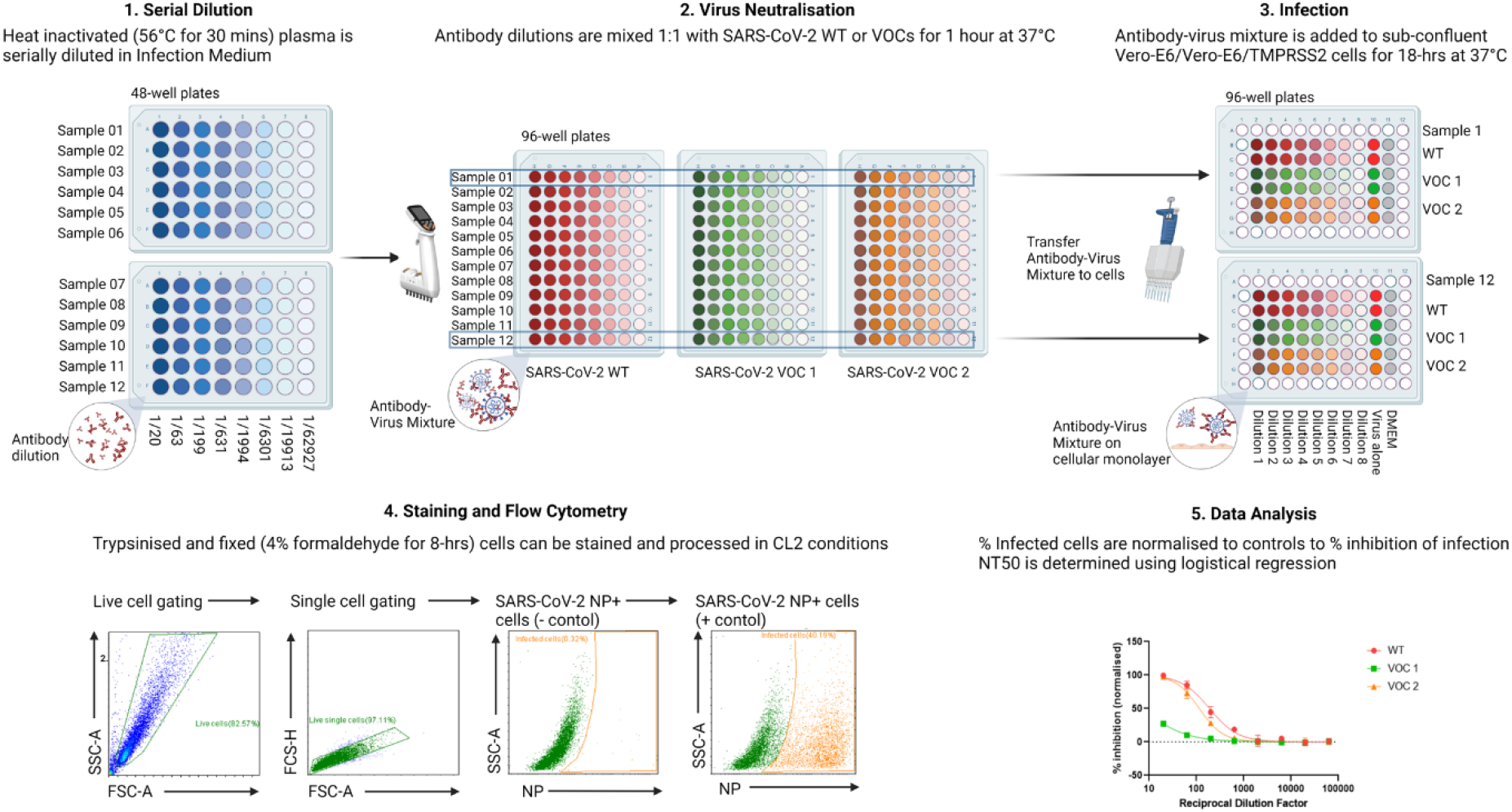
SARS-CoV-2 Flow Based Micro-Neutralisation Assay Workflow. 1. Heat inactivated (30 mins at 56°C) human plasma samples are serially diluted (8-point, half-log from 1/20) in Infection Medium, in 48-well plates. 2. Viral neutralisation is done in 96-well plates, where each antibody dilution is co-incubated with WT SARS-CoV-2 or a Variant of Concern (VOC) for 60 mins at 37°C. 3. Infection of sub-confluent Vero-E6 or Vero-E6/TMPRSS2 cells with antibody-virus mixture is carried out in 96-well plates for 18-hrs at 37°C. Each plate can test one plasma sample against 3 SARS-CoV-2 variants in parallel, in duplicates, including controls (Virus alone and Infection Medium alone). 4. Cells are trypsinised, fixed (4% formaldehyde for 8-hrs at room temperature), permeabilised and stained for intracellular SARS-CoV-2 Nucleoprotein (NP). Infection is measured using Flow Cytometry, first gating the live, single cell population, and then NP negative and positive populations. 5. Flow cytometry data is analysed to determine the % of infected (NP+) cells per well and then the % of inhibition can be determined by normalising each dilution to the positive and negative controls. NT50 can be determined from the inhibition curve, as the dilution factor resulting in 50% inhibition of infection. Created with BioRender.com

### Flow Cytometry-Based Micro-NT Workflow

Step 1: Plasma is heat inactivated for 30 mins at 37°C. This ensures inactivation of the complement system which can trigger cytolysis. For clinical trials, or when multiple experiments are planned per sample, after heat-inactivation the plasma can be aliquoted and stored at -80°C, to avoid multiple freeze-thaw events. Optional: prior to dilution, the samples can be filter-sterilised (0.25μM filter). If working with low volumes, this can be done after diluting the serum 1/20 in Infection Medium, and then proceeding with a half-log serial dilution (Figure 1). Minimum volume required is 10ul plasma for a test against a single variant, or 30ul against 3 variants (See Box 1 on sample processing).

BOX 1: Maximising Sample Processing

*Up to 12×96-well plates can be comfortably processed together where only the inner 60-wells of the plate are used, leaving the outer wells available to be filled with PBS, limiting evaporation from the wells, and avoiding the ‘edge-effect’. The 96-well plate format allows for three samples to be run in parallel, in duplicate, on a single plate (Figure 1), with a total of 36 samples per day, or a single sample against three SARS-CoV-2 variants for a total of 12 samples per day*.

*Within 1 hour, 12 plasma samples can be serially diluted across 2×48-well plates, using a 300ul multi-channel pipette, and divided amongst 3×96-well plates for incubation with SARS-CoV-2. An adjustable tip spacing, 300ul multi-channel makes it easy to transfer from the 48-well format to the 96-well format. The neutralisation takes one hour at 37°C 5% CO2. A second hour of hands-on time is then required to replace the medium on the cells from culture medium to infection medium (100ul per well) and add 100ul virus/plasma to the cells. As these are incubated in 96-well plates, this can easily be done with a standard or automated multi-channel pipette. This process takes 2-hrs for 12-plates*.

*Cells are trypsinised and transferred to round-bottom plates for fixation and staining. Transfer efficiency can be increased by using an electronic 1ml multi-channel pipette to serially dispense PBS/PFA across a full plate (Thermo Scientific #46300800). The percentage of infected cells is measured directly in 96-well plates through automatic flow-cytometric detection of intracellular SARS-CoV-2 N protein. Resuspending the cells in 60ul flow running buffer results in a run-time of 60-80 mins per plate, depending on instrument settings*.

Step 2: For viral neutralisation, plasma dilutions are co-incubated with an equal volume (1:1) live SARS-CoV-2 WT or VOCs for 1-hr at 37°C 5% CO_2,_ to facilitate antibody-virus interaction (Figure 1).

Step 3: Antibody-virus mixture is then added to sub-confluent Vero E6 or Vero E6/TMPRSS2 cells in 96-well plates and the cells are incubated for 18-hrs in 37°C, 5% CO_2_ (Fig 1.4). Controls include wells with no virus and virus alone. The SARS-CoV-2 variants used are fully infectious, capable of completing full replication and re-infection cycles, through cell-to-cell contact and also through release of infectious particles into the supernatant. 18-hrs is enough time for the cells to produce viral proteins abundantly, allowing for easy distinction of infected and non-infected cells during analysis, while not providing enough time for viral cytopathic effects to develop. This is important, as cells must be intact for subsequent staining and flow-cytometry analysis. See Box 2 for full details of the infectious model.

BOX 2: SARS-CoV-2 Infectious Model

*The assay was developed with Vero E6 cells or Vero E6/TMPRSS2 cells, as both express the ACE-2 receptor, are highly permissive to SARS-CoV-2, and are a standard cellular model for SARS-CoV-2 neutralisation assays (35-37). The cellular protease TMPRSS2 is a co-factor of SARS-CoV-2 entry expressed at the cell surface and important for membrane fusion. Its expression in Vero E6 enhances the rate of infection. Of note, amplification of SARS-CoV-2 on cells lacking the TMPRSS2 receptor can lead to a well-described cell-line adaption, whereby deletions or mutations in the furin-cleavage site, of 15-30bp at the S1/S2 cleavage site, allows the virus to become TMPRSS2-independent, in as few as two passages (38, 39). Furthermore, this deletion (ΔFurin) can be associated with reduced neutralisation compared to viruses bearing an intact Spike sequence, possibly due to an increased particle to PFU ratio, resulting in more particles per infectious dose (40)*.

*When multiple SARS-CoV-2 variants are tested in parallel, the stock viruses are titrated to infect 30-40% of cells after 18-hrs, as measured by flow cytometry. This provides a large enough population of infected cells to observe the neutralisation of the virus across the plasma dilution series. Infection of the population saturates around 70% infected cells, and so infection in this range could bias the resulting neutralisation titre*.

Step 4: At 18-hrs post-infection, and following trypsinisation, the cells are resuspended in PBS with a final concentration of 4% Formaldehyde to facilitate cell fixation and viral inactivation following an incubation of 8-hrs in the dark. Cells should be processed as soon as possible but can be stored in Formaldehyde for up to 24-hrs in the dark at room temperature. In instances where immediate processing is not possible, formaldehyde should be replaced with PBS and plates can be stored at 4°C in the dark for up to one week, although this is not recommended. Once the virus is inactivated, the plates can be transferred to Containment Level 2 facilities to be permeabilised and stained for intracellular SARS-CoV-2 nucleoprotein (NP). Flow cytometry is then used to measure NP positive cells, as a percentage of the total gated live, single-cell population for each well (Figure 1). The gating strategy is described in detail in Box 3.

BOX 3: Flow Cytometry Staining and Analysis Strategy

*Here we use intracellular SARS-CoV-2 N as our target protein. As N is more conserved than Spike (41), the same antibody can be used to detect WT-B, WT-B*.*177*.*18, Beta and Omicron-BA*.*5 variants. Furthermore, as N is the most abundantly expressed SARS-CoV-2 protein, it can be detected as soon as 6-hrs post-infection and is clearly distinguishable from uninfected cells by 18-hrs post-infection. Furthermore, there is no homology to cellular proteins that might result in high background staining*.

*Gating cells by Side Scatter (SSC) which indicates granularity, and Forward Scatter (FSC) which indicates size, allows viable cells to be distinguished from dead cells (low FSC, typical SSC) and cellular debris (low FSC and low SSC). Single cells can be separated from doublets or clumped cells by gating the FSC-Area against FSC-Height, which should be a linear relationship. We can then plot the live, single cells against FITC, which is the channel used to detect NP staining. Using our negative control wells (cells cultured in Infection Medium only, no virus) we gate to the immediate right of the cell population. As these cells should not contain any NP, <1% of cells should be considered positive. The same gate should then be applied to all wells. The positive controls (virus in Infection Medium, no plasma) should be checked to ensure there has been an appropriate level of infection*.

Step 5: Flow cytometry data can be exported as a CSV file for data analysis (Figure 1). Mean and standard deviation can be calculated for each well. Wells where there are <1000 live, single events do not have enough cells to be representative of the population and should be removed from the analysis. Background level in the negative control wells should be subtracted from all the readings. The data can then be presented as a percentage of inhibition of infection, where the negative controls have 100% inhibition, and the positive controls have 0% inhibition. The NT50 can then be determined by a four-parameter non-linear regression.

### Micro-NT produced comparable Neutralisation Titres to PRNT

We compared the NT50 values obtained using Micro-NT and conventional PRNT, as this is the gold standard viral neutralisation assay. We used WHO International Standard for anti-SARS-CoV-2 immunoglobulin (human), a pool of eleven human plasma samples from convalescent patients with Low, Medium, or High titre SARS-CoV-2 S IgG, intended as a calibrant for serological assays. The neutralising capacity of the standards against WT-B SARS-CoV-2 on Vero E6 cells was measured using conventional PRNT (Figure 2A) or Micro-NT (Figure 2B). Both assays were able to distinguish Low, Medium, and High IgG titre standards based on their neutralising capacity. The NT50 values were determined for each standard and compared between assays. The NT50 for the low IgG standard was very similar between the two assays 86 for PRNT, versus 62 with Micro-NT (Figure 2C). Both assays produced the same NT50 with the Medium IgG standard (435 with PRNT and 431 with Micro-NT). Both assays found a high NT50 with the High IgG standard, with Micro-NT showing greater separation between the Medium and High Titre results than the PRNT assay (1144 with PRNT and 2962 with Micro-NT). Interestingly, the fold change between the medium and high results with Micro-NT closely reflected the fold change between the antibody potencies in IU/ml as provided by the WHO (7-fold increase between medium and high IgGs using WHO NT50s and 6.9-fold increase using Micro-NT NT50s).

**Figure 2:**
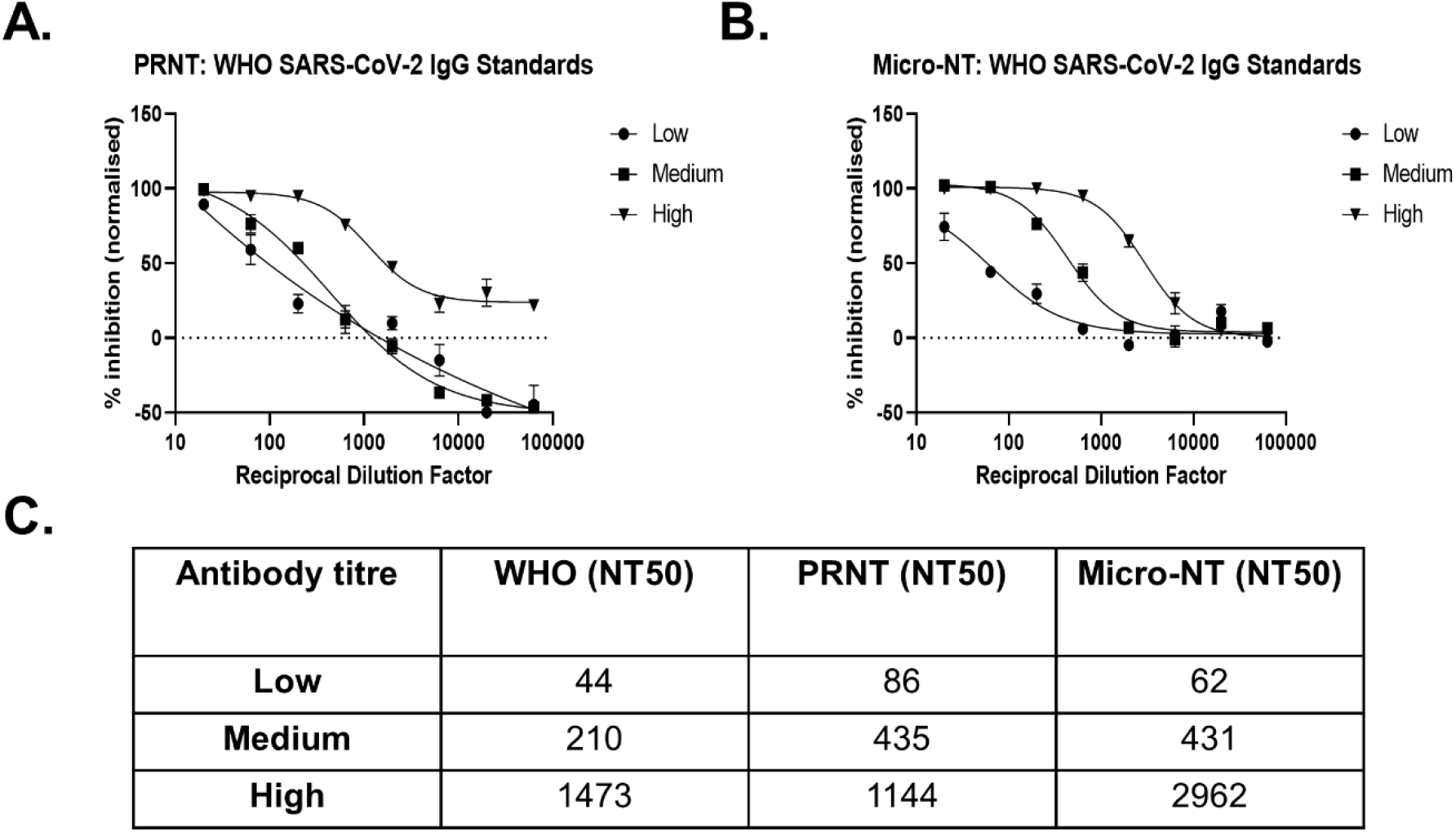
Validation of Flow-Based Micro-NT using WHO International SARS-CoV-2 IgG Standards (A-B) SARS-CoV-2 (WT-B) was neutralised for 1 hour with an 8-point, half-log serial dilution of WHO SARS-CoV-2 Low, Medium or High Titre IgG. (A) The antibody-virus mixture was then plated onto Vero E6 cells for 90 mins before overlay (1% CMC) was added and incubated for 96-hours. Plaques were quantified and the inhibition of infection is presented as a percentage, relative to the virus-only control wells. Graph shows the mean and standard deviation of duplicates (n=1). (B) In parallel, the virus-antibody mixture was added onto Vero E6 cells for 18-hrs before cells were trypsinised, fixed and permeabilised, and stained for the SARS-CoV-2 NP. % of infected cells was determined by flow cytometry analysis. Inhibition of infection is presented as percentage, relative to the virus-only control wells. Graph shows the mean and standard deviation of duplicates (n=1). (C) Table shows the potency of Neutralising antibodies present in WHO International SARS-CoV-2 IgG Standards with Low, Medium and High titre IgG in International Units (IU/ml) as determined by the WHO. NT50 values were determined for each standard by running in parallel on PRNT and Micro-NT, against SARS-CoV-2 (WT-B) on Vero E6 cells. NT50 was determined using the percentage of inhibition for each dilution factor, relative to the virus-only control and calculated from a non-linear regression (variable slope) with Graph Pad Prism equal to the 50% Inhibitory Dilution (ID50).

### Micro-NT has good inter-assay and intra-assay reproducibility

The Flow-Cytometry based Micro-NT assay reproducibility was determined by measuring Inter-Assay and Intra-assay variability. To determine the Intra-assay reproducibility, we ran 3 COVID-19 convalescent plasma samples, with a low, medium or high NT50, each with an 8-point, half-log serial dilution (Figure 3A-3C). Each dilution was tested in 6 replicate wells on the same plate and the mean and standard deviation calculated. The mean standard deviation was 1.76% across all 3 plates, with a range from 0.09% (mean infection 0.37%) to 3.57% (mean infection 49.2%), showing high intra-assay reproducibility.

**Figure 3:**
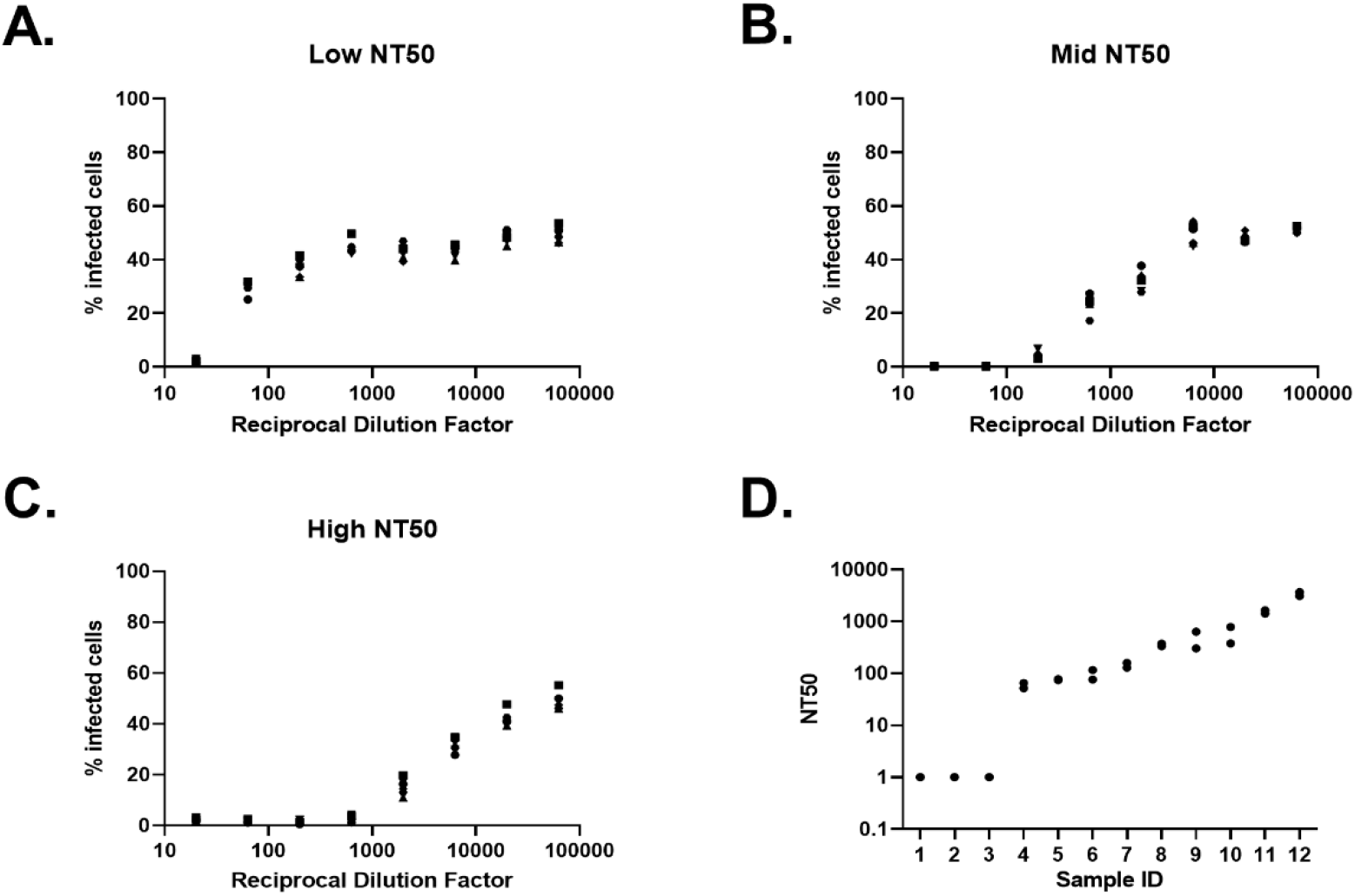
Flow Cytometry-Based Micro-NT Intra- and Inter-Assay Variation (A-C) Data shows the % infection across 8 dilutions of convalescent sera with (A) low, (B) mid, or (C) high neutralising capacity against WT-B.177.18 SARS-CoV-2 on Vero E6/TMPRSS2 cells. Each dilution was tested in 6 wells on a single plate, and the % infected cells in each well is shown. (D) NT50 values for 12 convalescent serum samples against WT-B.177.18 SARS-CoV-2 were analysed using Micro-NT. Each dilution was tested in duplicate, and each sample was run twice, independently. NT50 was determined using the percentage of inhibition for each dilution factor, relative to the virus-only control and calculated from a non-linear regression (variable slope) with Graph Pad Prism equal to the 50% Inhibitory Dilution (ID50). Graph shows the NT50 values obtained for each sample (n=2).

3 naïve and 9 COVID-19 convalescent plasma samples were analysed using Micro-NT to determine the NT50 (Figure 3D). Each sample was tested using technical duplicates, in two independent experiments. The NT50 was determined for each experiment. The Mean and Standard Deviation was determined for each sample, and the co-efficient of variation (CV) measured (Standard Deviation/Mean). The mean CV was 11.64%, which shows high inter-assay reproducibility (Supplementary Table 1). The naive samples displayed no neutralising capacity (NT50<20).

### Micro-NT has a broad Dynamic Range

Across 190 convalescent plasma samples tested, we have found a broad range distribution of neutralising capacities, from 1 to >5000, without saturating the assay. The lower Limit of Detection (LOD) of the assay is an NT50 of 20, corresponding to the initial antibody dilution. Any sample unable to achieve a minimum of 50% inhibition with the initial dilution is given an arbitrary NT50 of 1. An 8-point, half-log dilution from 1/20, to 1/62927, provides an optimal range for measuring neutralising capacity against SARS-CoV-2 from convalescent (Figure 4A-4D) or vaccinated (data not shown) plasma samples.

**Figure 4:**
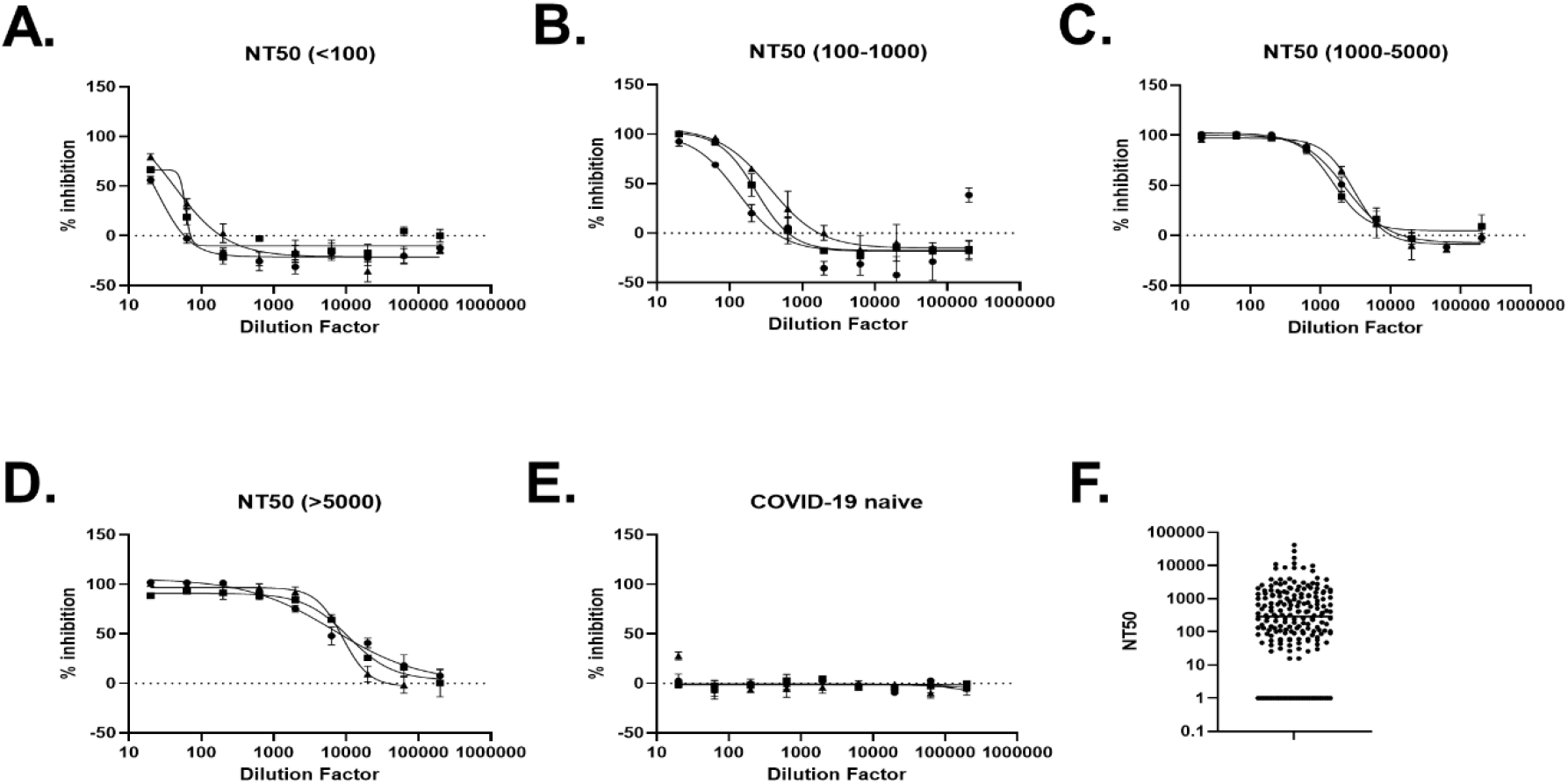
Dynamic Range of Flow Cytometry-Based Micro-NT We analysed the NT50s of 190 COVID-19 naïve or convalescent plasma samples, using Micro-NT, against SARS-CoV-2 (WT-B) on Vero E6 for 18-hrs. (A-D) Graphs show neutralising capacity of 3 convalescent plasma samples (P1-P3) across 8-point, half-log serial dilution, representative of samples with 50% Neutralisation Titres (NT50) within a similar range. The percentage of viral inhibition of each dilution was calculated relative to the virus only control. (E) Plasma from COVID-19 naïve individuals showed little or no neutralisation capacity against SARS-CoV-2. (F) Graph shows all the NT50 values obtained across the cohort of COVID-19 convalescent plasma samples, measured using Micro-NT.

### Micro-NT can be used to Measure Modulation in Neutralising Capacity against SARS-CoV-2 Variants of Concern

To examine whether Micro-NT can identify and characterise immune-escape SARS-CoV-2 VOCs, we used the WHO SARS-CoV-2 IgG Standards in parallel to neutralise WT-B.177.18 SARS-CoV-2, and two VOCs, Beta and Omicron-BA.5 on Vero E6/TMPRSS2 cells (Figure 5A-C). When neutralised with Low IgG, WT-B.177.18 was the only variant neutralised (NT50 of 98.72), while neutralisation of Beta and Omicron-BA.5 was undetectable (Figure 5D). With Medium Titre IgG, all variants were neutralised (NT50s of 192.1, 31.94 and 31.38 respectively), though with a 6-fold decrease in capacity against Beta and Omicron-BA.5 compared to WT-B.177.18. With High Titre IgG, WT-B.177.18 remained the most effectively neutralised, with an NT50 of 919.7, while Beta and Omicron-BA.5 remained poorly neutralised with a 4.5-fold decrease seen with Beta (NT50 of 203.4) and a 7.3-fold decrease seen with Omicron-BA.5 (NT50 of 125.4).

**Figure 5:**
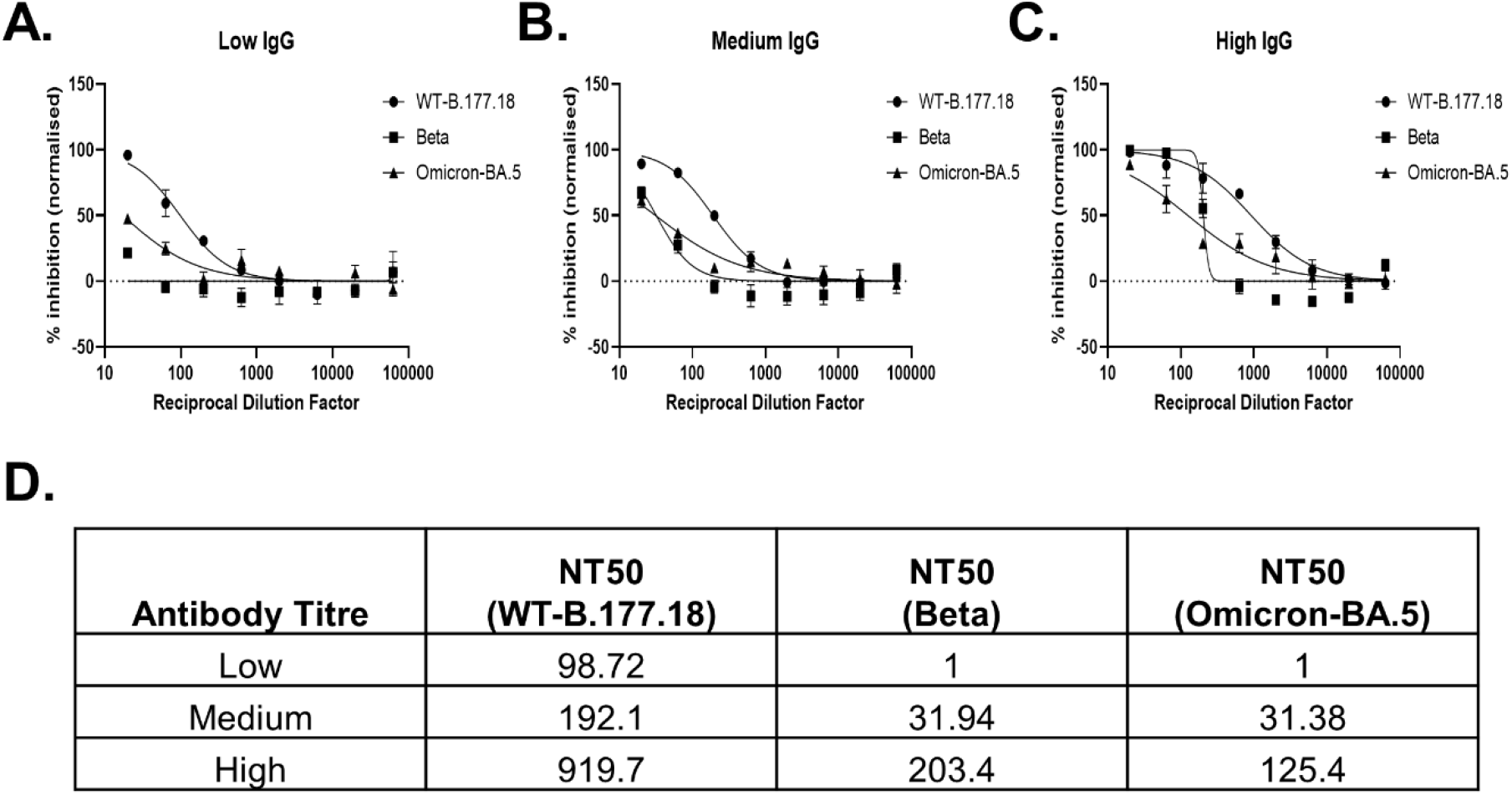
Comparison of SARS-CoV-2 IgG Standards tested against SARS-CoV-2 Variants SARS-CoV-2 WT-B.177.18, Beta and Omicron-BA.5 were neutralised for 1 hour with an 8-point, half-log serial dilution of WHO SARS-CoV-2 (A) Low, (B) Medium or (C) High Titre IgG. The viruses were plated onto Vero E6/TMPRSS2 cells for 18-hrs before cells were trypsinised, fixed and permeabilised, and stained for the SARS-CoV-2 NP. % of infected cells was determined by flow cytometry analysis. Inhibition of infection is presented as percentage, relative to the virus-only control wells. Graph shows the mean and standard deviation of duplicates (n=1). (D) shows the NT50 values obtained for the WHO International SARS-CoV-2 IgG Standards with Low, Medium and High titre IgG when run against WT-B.177.18 SARS-CoV-2, and VOCs Beta and Omicron-BA.5. NT50 was determined using the percentage of inhibition for each dilution factor, relative to the virus-only control and calculated from a non-linear regression (variable slope) with Graph Pad Prism equal to the 50% Inhibitory Dilution (ID50).

This flow-cytometry based Micro-NT is a medium throughput, reproducible, live SARS-CoV-2 assay, comparable to PRNT but suitable for screening large clinical cohorts for NAbs against SARS-CoV-2 variants

## Conclusion

Here we have introduced and presented in detail a new flow-cytometry-based Micro-Neutralisation Assay to quantify the neutralisation capacity of antibodies from plasma or serum. Live SARS-CoV-2 is neutralised by antibodies from convalescent/vaccinated plasma and then used to infect Vero-E6/Vero-E6/TMPRSS2 cells in a 96-well plate format. After 18-hrs, cells are trypsinised, fixed and stained for intra-cellular SARS-CoV-2 NP. Flow cytometry is then used to quantify the % of infected cells per condition, which allows determination of the NT50 for each plasma sample, against WT SARS-CoV-2 or VOCs.

This approach has been calibrated and validated against WHO SARS-CoV-2 IgG standards, using WT and VOCs. Using WT-B SARS-CoV-2, we have shown comparable NT50 titres obtained using PRNT, the gold standard for serological testing, and our flow-based Micro-NT, with the advantage of significantly increased throughput. This assay is medium throughput, capable of processing 3 samples per 96-well plate, with 12 plates being comfortably processed per day, simultaneously, for a total of 36-samples per day. The 18-hr infection duration is at the lowest end of the requirements for other micro-NTs (Table 1). Unlike CPE-based assays it directly measures infection through the detection of SARS-CoV-2 NP in infected cells, minimising risks of contamination or toxicity from serum/plasma being interpreted as viral-CPE. Similar to using well imagers to measure spot-forming units, flow cytometry quantifies the infection in each well, meaning a dose-response curve can be plotted allowing determination of the NT50.

Flow cytometry has several advantages as an assay endpoint. Firstly, infection is quantifiable, being able to see the number of infected cells as well as the percentage of the whole population. This is in contrast to a technique such as RT-qPCR where normalisation against a house-keeping gene is required to interpret the data, or a CPE-based approach, where wells can be scored as positive or negative, but not on a gradient. Secondly, the size and granularity of the cell population is measured, allowing issues with viability or contamination to be quickly identified, that may not have been visible with a light microscope. Thirdly, a flow cytometer with an automatic plate reader will process all wells without requiring user input, other than set-up and cleaning, limiting hands-on processing time. Furthermore, as cells are fixed prior to staining, flow analysis can take place in CL2 conditions, when compact flow cytometers are placed within Biosafety Cabinets to control for risk of aerosol formation. This protocol expands the option to perform live cell neutralising assays to those with flow-cytometry facilities, which is becoming more common as labs are investing in this popular and diversely applicable technique. However, as with all live virus assays, limitations include access to CL3 facilities, with trained staff to carry out live virus work.

While pseudovirus assays performed in CL2 conditions can address some of these limitations, live-virus assays remain the gold standard. New VOCs can be rapidly isolated from nasopharyngeal swabs, and once validated, can be used directly in the assay. By titrating the viruses to infect the same % of cells after 18-hrs, neutralisation capacity can be determined against several SARS-CoV-2 variants in parallel, without needing to account for differences in CPE, or duration of time to development of CPE. The assay is also appropriate for testing antibodies from a variety of sources, including serum, plasma, or commercial monoclonal antibodies.

Flexibility in SARS-CoV-2 variants and antibody sources means the assay is easily adapted to multiple applications. We have used the flow-based Micro-NT to define an RBD titre post-infection/vaccination that correlates with a protective neutralisation titre (Kenny et al, in review) as well as to identify COVID-19 convalescent individuals as donors for convalescent plasma therapy. Another use will be measuring functional antibody production post-vaccination. During initial COVID-19 vaccine clinical trials, high infection and hospitalisation rates allowed for collection of data on protection from infection or severe disease (42-45). Indeed, neutralisation titres post vaccination were found to be directly correlated with vaccine efficacy (46). However, as vaccine uptake increases, and COVID-19 cases decrease, such clinical endpoints are harder to achieve. Surrogate endpoints, including neutralization titres will therefore play an important role in measuring vaccine efficacy in next generation vaccine trials (46, 47). VACCELERATE, an EU Consortium is currently co-ordinating clinical trials identifying vaccine efficacy in elderly (ClinicalTrials.gov Identifier: NCT05160766) and paediatric populations (EudraCT Number: 2021-005043-71) as well as the safety, timing, and efficacy of COVID-19 booster vaccines in the fully vaccinated (EudraCT number 2021-004889-35). The flow-cytometry based micro-neutralisation assay described here was chosen to enable the measurement of NAbs across large numbers of clinical specimens (>400) across multiple SARS-CoV-2 variants.

## Methods

### Cells

Vero E6 (VERO C1008, Vero 76, clone E6, Vero E6), were obtained from ATCC (ATCC CRL-1587) and maintained in Dulbecco’s Modified Eagle’s Medium (DMEM, Thermo Scientific, 61965-026) supplemented with 10% Foetal Bovine Serum (Themo Scientific, 10500-064) (DMEM-10). Vero E6/TMPRSS2 cells (#100978), obtained from the Centre For AIDS Reagents (CFAR) at the National Institute for Biological Standards and Control (NIBSC) (48) were cultured in DMEM supplemented with 10% Foetal Bovine Serum (FBS, DMEM-10) and Geneticin (Thermo Scientific, 10131035) at a concentration of 1mg/ml, at 37°C in 5% CO2. All cell lines routinely tested negative for mycoplasma. During the course of infection, cells were incubated in Complete Infection Medium, DMEM 2% FCS (DMEM-2) supplemented with Penicillin (1U/ml) and Streptomycin (100ug/ml) (Sigma-Aldrich, P4333) and amphotericin B (0.5ug/ml) (Thermo Fisher Scientific, 15290018)

### Human Plasma samples

Plasma derived from ethylenediamintetraacetic acid (EDTA) anticoagulated whole blood was collected from COVID-19 RT-qPCR positive and negative participants of the All-Ireland Infectious Disease (AIID) Cohort (49). For this analysis we included either individuals with polymerase chain reaction (PCR)-confirmed COVID-19, documented vaccination with 2 doses of mRNA-1273, BNT162b2 or ChAdOx1-S, at least 14 days from the second dose, or individuals without COVID-19 or vaccination recruited with available bio-banked plasma dated no later than July 2019, prior to the onset of the COVID-19 pandemic. A total of 190 convalescent plasma samples from COVID-19 patients were tested in this study. 11 non-COVID-19 plasma samples, collected prior to the COVID-19 pandemic and negative for anti-SARS-CoV-2 antibodies were included as negative controls. Samples were stored at -80°C before use. All human plasma samples were heat-inactivated at 56°C for 30 minutes before testing. First WHO International Standard for anti-SARS-CoV-2 immunoglobulin, human (NIBSC code: 20/136) is a pool of eleven human plasma from convalescent patients and was established in December 2020 by the WHO Expert Committee on Biological Standardization.

### SARS-CoV-2 clinical isolates

All work with live SARS-CoV-2 was carried out in Containment Level 3 laboratory under Biosafety Level 3 guidelines. WT Spike, Pango lineage B (WT-B) SARS-CoV-2 clinical isolate 2019-nCoV/Italy-INMI1, Clade V, Passage 4 (50) was obtained from the European Virus Archive goes Global (EVAg, Spallanzani Institute, Rome) and following its amplification on Vero E6 cells (Passage 6) contained a deletion in the Furin cleavage site. WT SARS-CoV-2 with D614G substitution (Pango lineage B.177.18 (WT-B.177.18), GenBank accession ON350866, Passage 2), Beta (Pango lineage B.1.351, GenBank accession ON350868, Passage 2) and Omicron (Pango lineage BA.5 (Omicron-BA.5), GenBank accession OP508004, Passage 1) clinical isolates were isolated from SARS-CoV-2 positive nasopharyngeal swabs from the AIID cohort. Supernatant was filter sterilised (0.2nm filter) and diluted 1:1 with Complete Infection Medium. Confluent Vero E6/TMPRSS2 cells were incubated with the virus in a T12.5 flask until 50% cell death was observed using a light microscope. The supernatant and cells were transferred to a confluent T175, with Complete Infection Medium. Cells were cultured until 50% cell death was observed. The supernatant was collected and centrifuged at 4000xg for 5 minutes, aliquoted and stored at -80°C. The titre of the virus stocks was determined by median Tissue Culture Infectious Dose assay (TCID50) and/or plaque assay. Viral RNA was isolated using Qiagen Viral mRNA mini-kit according to manufacturer instructions. Viral RNA genome was sequenced to confirm the integrity of the Spike protein in each variant and lineage. Viral genomes were quantified by RT-qPCR amplification of the SARS-CoV-2 Nucleoprotein (N2) gene (RNA diluted 1 in 10 in nuclease-free water resulted in Ct values of 19.5, 21.1 and 20.1 for WT-B.177.18, Beta and Omicron-BA.5 variants, respectively. WT-B.177.18 had a titre of 2.6×10^6^ PFU per ml, Beta had a titre of 1.2×10^6^ PFU/ml and Omicron-BA.5 had a titre of 2.2×10^6^ PFU/ml.

### Flow Cytometry-Based Micro-Neutralisation Assay

#### Viral Neutralisation

Heat-inactivated plasma samples were diluted with a half-log, 8-point serial dilution, from a starting dilution of 1 in 20 in Complete Infection Medium. SARS-CoV-2 isolates were titrated to infect 30-40% of Vero E6/Vero E6/TMPRSS2 cells 18-hr post infection as determined by flow cytometry analyses of SARS-CoV-2 Nucleoprotein positive cells. Viral isolates were diluted in Complete Infection Medium and co-incubated with the plasma dilutions in a 1:1 ratio for 1 hour at 37°C, 5% CO_2._

#### Infection

Vero E6 or Vero E6/TMPRSS2 cells were plated (2.5×10^4^) the day before use in 100ul Culture Medium, in clear flat-bottom 96-well plates (Sarstedt, 83.3924). The cells were incubated overnight at 37°C 5% CO_2_ to reach 90-100% confluency at time of infection. Post-neutralisation, supernatant was removed from the cells and replaced with 100ul Infection Medium and 100ul virus/plasma mix, in triplicates or duplicates. Positive controls (Virus alone) and Negative controls (Infection Medium alone) were included in each plate. Cells were incubated for 18 hours at 37°C 5% CO_2_.

#### Cell collection

Supernatant was discarded and cells were rinsed with 100ul PBS. 25ul Trypsin-EDTA (Gibco, # 25300054) was added to the cells at 37°C until single-cell suspension was obtained. Cells were resuspended in 75ul PBS and transferred to a round-bottom 96-well plate (Sarstedt, 83.3925500) with 100ul 8% formaldehyde solution (Sigma Aldrich, F8775) to achieve a final concentration of 4%. Cells were fixed in the dark at room temperature for a minimum of 8 hours.

#### Cell Staining

All steps-post fixation were carried out in Biosafety Level 2 laboratory in Class 2 Biosafety Cabinets. All centrifugation was carried out at 4000xg for 5 minutes (Heraeus Megafuge 16R, Thermo Scientific), with 96-well plates contained within sealed buckets (Thermo Scientific, #75003625). The supernatant was removed, and cells were permeabilised with Perm/Wash Buffer (BD, 554723) according to manufacturer’s instructions, which was maintained throughout antibody staining. Intracellular SARS-CoV-2 Nucleoprotein (NP) staining was performed with SARS/SARS-CoV-2 Nucleocapsid Monoclonal Antibody (E16C) (1/100 dilution, Invitrogen, MA1-7403), goat anti-mouse IgG2b-FITC (1/500 dilution, Santa Cruz Biotechnology, SC-2080).

#### Flow Cytometry

Cells were resuspended in 60ul PBS-EDTA-2% for flow cytometry analysis (Beckman Coulter CytoFlex or CytoFlex S). Forward and Side-Scatter gates were used to exclude debris from intact cells. Cells were then gated using Forward Scatter Area where single cells were gated based on linearity between Area and Height, excluding doublets. % Infected cells of the single cell population were determined using the negative control wells to set the boundary of the negatively staining populating in the FITC-channel (Blue 488nm laser, 525/40 filter). Gating was performed using CytExpert software (version 2.4.0.28, Beckman Coulter).

#### Analysis

Wells with <1000 single events were excluded from analysis. Positive controls (virus only, no plasma) had to be >20% and <70% infected while negative controls (Infection Medium only) had to be <3% or the plate would be excluded from analysis. The mean % infected cells for each plasma dilution was calculated, and this result was normalised using the control wells, where 0% viral inhibition was equal to the positive control and 100% viral inhibition was equal to the negative control. The plasma dilution resulting in a 50% reduction in infection (NT50) was determined using logistical regression (4-paramater, variable slope) with GraphPad Prism (Version 9.3.1).

### Plaque Reduction Neutralisation Test (PRNT)

Vero E6 cells were plated (4.2×10^5^) in 1ml DMEM-10 in clear flat-bottom 12-well plates. The cells were incubated overnight at 37°C 5% CO_2_ to reach 100% confluency at time of infection. WHO SARS-CoV-2 IgG Standards with Low, Medium and High Titre IgG were diluted with a half-log, 8-point serial dilution, from a starting dilution of 1 in 20 in Compete Infection Medium. SARS-CoV-2 (WT-B) was diluted in Complete Infection Medium to a concentration of 100 Plaque Forming Units (PFU) per 100ul and co-incubated with the WHO standard dilutions in a 1:1 ratio for 1 hour at 37°C 5% CO_2_. Supernatant was removed from the cells and 200ul plasma/virus was added to each well. Plate was gently shaken to ensure the surface of the well was evenly coated. The plates were incubated at 37°C 5% CO_2_ for 90 mins, with shaking every 10 mins. 2ml Complete Infection Medium with 1% carboxycellulose (CMC, Sigma #C5013) was added to each well. The plates were incubated for 96-hours until plaques became visible. 2ml 8% formaldehyde solution was added directly to each well to a final concentration of 4% and the plates were fixed for a minimum of 8 hours in the dark at room temperature. The supernatant was removed from the cells and the cells washed 3 times in ddH_2_O until all CMC was removed. Crystal violet (Sigma-Aldrich, #HT90132) 0.5% was added to just cover the surface of the well, incubated for 15 mins at room temperature, then removed with 2 washes in ddH_2_O. Plates were allowed to fully dry before being photographed. A modified ImageJ script (51) was used to count the plaques per well and the duplicates were averaged and normalised to the positive control (virus only). The % inhibition per dilution was analysed with a non-linear regression (variable slope) on GraphPad Prism to determine the NT50.

## Data Availability

All data produced in the present work are contained in the manuscript

## Abbreviations

S: Spike
NP: Nucleoprotein
NAbs: Neutralising Antibodies
RBD: Receptor Binding Domain
ACE-2: Angiotensin-Converting Enzyme 2
WT: Wild-Type
VOC: Variant of Concern
PRNT: Plaque Reduction Neutralisation Test
CPE: Cytopathic Effects
NT50: 50% Neutralisation Titre.

## Acknowledgements

SOR is the recipient of the Irish Research Council (IRC) Government of Ireland Postgraduate Scholarship (GOIPG/2019/4432). This study was supported by Science Foundation Ireland (20/COV/0305) and VACCELERATE, the European Corona Vaccine Trial Accelerator Platform. GK is funded through a fellowship from the United States Embassy in Ireland. The authors wish to thank all study participants and their families for their participation and support in the conduct of the All-Ireland Infectious Diseases Cohort Study.

The work has been performed with contribution or support of VACCELERATE participants.

The All-Ireland Infectious Diseases Cohort Study Investigators: Mater Misericordiae University Hospital: A. Cotter, E. Muldoon, G. Sheehan, T. McGinty, JS. Lambert, S. Green, K. Leamy. St Vincent’s University Hospital: G. Kenny, K. McCann, R. McCann, C. O’Broin, S. Waqas, S. Savinelli, E. Feeney, PWG. Mallon. CEPHR: A. Garcia Leon, S. Miles, D. Alalwan, R. Negi. Beaumont Hospital: E. de Barra, S. McConkey, K. Hurley, I. Sulaiman. University College Cork: M. Horgan, C. Sadlier, J. Eustace. University College Dublin: C. Kelly, T. Bracken. Sligo University Hospital: B. Whelan, Our Lady of Lourdes Hospital: J Low. Wexford General Hospital: O Yousif. University Hospital Galway: B. McNicholas. St Luke’s Hospital Kilkenny: G. Courtney. Children’s Health Ireland: P. Gavin.

---

**Supplementary Table 1:**
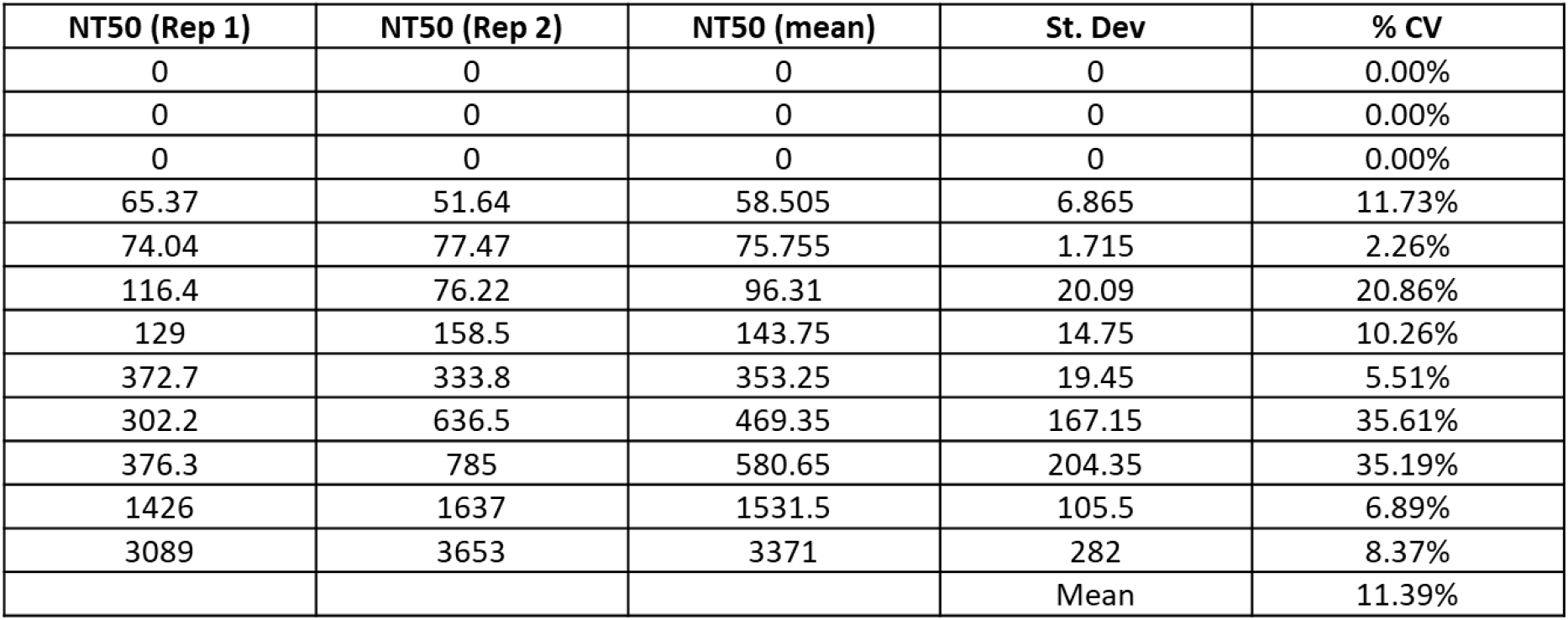
Inter-Assay Co-efficient of Variation Table shows the NT50 values obtained from 2 independent experiments of 12 convalescent COVID-19 plasma samples, as measured using Micro-NT against D614G SARS-CoV-2. Co-efficient of Variation (CV) was calculated as Standard Deviation (St. Dev)/Mean of Replicate 1 and 2 and displayed as a percentage. The % CV between plates is the average CV of all samples tested.

